# Four-Year Evaluation of a Pharmacists-Providers Collaborative Iron Deficiency Clinic in Heart Failure Care

**DOI:** 10.64898/2026.01.05.26343488

**Authors:** Kazuhiko Kido, Christopher Bianco, Marco Caccamo, Brittany Carey, Bailey Colvin, Timothy Dotson, Kevin Felpel, Lindsay Hedrick, Olivia Lackovic, Cindy McMahon, Rachael Schauble, Monica Thomas, George Sokos

**Affiliations:** Department of Clinical Pharmacy, West Virginia University School of Pharmacy, Morgantown, WV, U.S.A; Department of Medicine, West Virginia University Medicine, Morgantown, WV, U.S.A; Heart Vascular Institute, West Virginia University Medicine, Morgantown, WV, U.S.A; Department of Pharmacy, West Virginia University Medicine, Morgantown, WV, U.S.A; West Virginia Clinical and Translational Science Institute, Morgantown, WV, U.S.A; Division of Cardiovascular Medicine, University of Utah, Salt Lake City, UT, U.S.A; West Virginia University School of Pharmacy, Morgantown, WV, U.S.A.

**Keywords:** iron, heart failure, iron deficiency, multidisciplinary, pharmacist, quality of life

## Abstract

**Purpose:** To evaluate the performance of the pharmacists-providers collaborative iron deficiency treatment clinic in the heart failure service

**Methods:** A single-center retrospective cohort study was conducted to evaluate the performance of the iron deficiency pharmacists-providers collaborative care clinic during the induction and maintenance phases of intravenous (IV) iron therapy. The study included patients who were seen by HF providers (advanced HF cardiologists or advanced practice providers) and received the IV iron consultation with HF pharmacists. The study included patients aged 18 years or older who were diagnosed with HF or PH and received at least one dose of either ferric carboxymaltose or iron sucrose in outpatient settings. It was managed by the pharmacists-providers collaborative IV iron clinic. The primary outcome was adherence to the IV iron appropriate use criteria, laboratory requirements, and dosing during the induction course. The primary and secondary outcomes were compared with those of the previously reported control group, which received the usual iron deficiency treatment care at the HF clinic prior to the implementation of the pharmacists-providers collaborative IV iron deficiency treatment clinic. The use of oral iron therapy was evaluated over four years.

**Results:** A total of 187 patients were included in the final cohort. The median follow-up period of the IV iron consulting team was 372 (176, 623) days. Compared to the pre-implementation group, the primary outcome was significantly higher in the post-implementation group (81.3 vs. 40%, p<0.001). The most common reasons for nonadherence were the absence of maintenance laboratory requirements (15.5%), failure to administer all induction doses (1.6%), inappropriate use (1.1%), and incorrect dose (0.5%). Ferritin, iron saturation, and hemoglobin values at cycle one were significantly increased after the IV iron induction course compared to baseline values and remained numerically stable throughout the follow-up periods. Serum phosphorus levels remained within the normal range throughout the follow-up period. Among 57 patients on oral iron therapy, the consulting team discontinued it in 19 patients (33.3%) during follow-up.

**Conclusion:** The pharmacists-providers collaborative IV iron clinic significantly improved the performance of IV iron therapy in rural heart failure care settings compared to the usual care. These results highlight the importance of multidisciplinary care management for iron deficiency in the real-world heart failure practice.

## Introduction

Iron deficiency in heart failure (HF) occurs in more than 50% of patients with HF.^1^ Iron deficiency is significantly associated with worse functionality, lower quality of life, and worse mortality.^2^ Multiple clinical trials have shown that intravenous (IV) iron therapy improved functionality and quality of life and reduced hospitalizations for HF.^3–6^ The American College of Cardiology/American Heart Association/Heart Failure Society of America HF guidelines recommend IV iron therapy for patients with HF with left ventricular ejection fraction < 45 %.^7^ However, only one-fifth of patients meeting the criteria for IV iron therapy received IV iron in the real-world practice, potentially due to a lack of implementation strategies for IV iron therapy, access to IV iron infusion centers, care coordination issues, and time constraints of HF providers.^8,9^ Authors previously documented the first reported pharmacists-providers collaborative iron deficiency treatment clinic to overcome these implementation barriers.^10^ The initial pilot study showed significant improvements in the quality of iron deficiency care and doubled the number of patients who received IV iron therapy during the induction course. However, the initial report described only the clinic’s short-term performance during the induction course, and the number of patients was small. The primary objective of the study was to evaluate the 4-year-long-term experiences of this pharmacists-providers collaborative iron deficiency clinic during both the induction and maintenance phases in a larger-scale cohort.

## Methods

### Description of the clinical service

Authors previously reported details of this multidisciplinary pharmacists-providers collaborative iron deficiency service.^10^ Briefly, the HF services offer a pharmacists-providers collaborative iron deficiency care service mainly for outpatients, and it recently initiated a transition-of-care service for IV iron to improve care transitions from inpatient to outpatient settings. Once HF providers (advanced HF cardiologists or advanced practice providers) identify candidates for IV iron therapy after screening for iron deficiency, HF pharmacists received consulting orders from HF providers and lead the entire process, including IV iron therapy eligibility, development of an IV iron therapy plan, referrals to local infusion centers based on the patient’s residential location, and monitoring recipients of IV iron therapy. The referrals were placed both in outpatient and inpatient settings. Once patients receive an IV iron induction course, iron studies and complete blood counts (CBCs) were obtained 3 to 6 months after the induction course ends. HF pharmacists assessed the need for IV iron maintenance therapy and develop it longitudinally. Ferric carboxymaltose was the preferred agent, and iron sucrose was the second-line agent in outpatient settings, depending on the insurance formularies and local infusion center availability. If ferric carboxymaltose was selected, phosphorus is measured regularly. Insurance specialists authorized all IV iron therapy plans through insurance, if required.

### Study design

A single-center retrospective cohort study was conducted to evaluate the performance of the iron deficiency pharmacists-providers collaborative care clinic during the induction and maintenance phases. The study included patients who were seen by HF providers and received an IV iron consultation with HF pharmacists from January 1, 2021, to May 30, 2025. The study included patients aged 18 years or older who were diagnosed with HF or PH, completed the induction course of either ferric carboxymaltose or iron sucrose in an outpatient setting, and were managed by the pharmacist-provider collaborative IV iron clinic. Iron deficiency was defined as either ferritin < 100 mcg/L or ferritin level 100-299 mcg/L and iron saturation < 20%. Hemoglobin levels greater than 15 mg/dL were excluded. Patients who received the initial doses of IV iron prior to 2021 were excluded from the primary analysis but included in the cohort when the number of IV iron doses coordinated by the HF pharmacists was summarized. The primary outcome was adherence to the IV iron appropriate use criteria, laboratory requirements, and dosing during the induction course (Table 1). The appropriate criteria were developed based on the HFSA statement, HF guidelines, and the published expert consensus.^1,7,9^ Reasons for non-adherence were selected from the options: no maintenance iron study or hemoglobin levels within 6 months after the induction course, incorrect dose, and inappropriate use. The secondary outcomes included 1-year hospitalization for HF, all-cause mortality, EF within 1 year after induction course therapy, and discontinuation of oral iron therapy. Longitudinal changes in iron study laboratory results and the use of oral iron therapy were evaluated over four years. Adverse events were also reported.

**Table 1.** Appropriate criteria for intravenous iron therapy use, dose and laboratory monitoring.

|  |  | Weight <70 kg |  |  | Weight ≥ 70 kg |  |  | Infusion instructions |
| --- | --- | --- | --- | --- | --- | --- | --- | --- |
|  | Hgb | < 10 g/dL | 10-14 g/dL | 14 < to < 15 g/dL | < 10 g/dL | 10-14 g/dL | 14 < to < 15 g/dL | Undiluted bolus injection with 100 mg/min or 15 minutes for 1000 mg |
| Induction course | Day 0 | 750-1000 mg | 750-1000 mg | 500 mg | 750-1000 mg | 750 mg | 500 mg |  |
|  | Week 1-6 | 500 mg | No dose | No dose | 750-1000 mg | 750 mg | No dose |  |
| Maintenance course | Every 3-6 months | 500 mg if ferritin < 100 ng/mL or 100-300 ng/mL with Tsat <20% |  |  |  |  |  |  |

The primary and some key secondary outcomes were compared with those of the previously reported control group, which received usual iron deficiency treatment at the HF clinic prior to the implementation of the pharmacists-providers collaborative IV iron deficiency treatment clinic. The distribution of IV iron therapy administered throughout the Appalachian region was visually described. The rural area was defined with the Health Resources and Services Administration Rural Health Grants Eligibility Analyzer.^11^ The effect of the estimated distances between patients’ residential locations and infusion centers on the outcomes was also evaluated as the potential reason for nonadherence to the recommended IV iron therapy. For geospatial analysis, patients’ zip codes and infusion centers’ zip codes were geocoded to their respective zip code centroids. Drive times for zip codes were determined by the location of their centroids within a drive-time band. A rural drive-time analysis was performed by creating rural drive-time bands using ESRI ArcOnline Map’s Generate Travel Areas tool in fifteen-minute intervals up to seventy-five minutes.^12^ A rural drive time was also created for WVU Medicine Ruby Memorial Hospital, where the HF clinic is located, in fifteen-minute intervals up to five hours, where was the main infusion center for IV iron therapy prior to the implementation of this multidisciplinary pharmacists-providers collaborative iron deficiency service. A density heat map was created to show where the raw numbers of patients were distributed throughout the study area. Maps were created for both rural drive times for comparison. The Ruby rural drive time bands were displayed in hour intervals for clarity. A bivariate map was created comparing rural drive time to an infusion center versus the IV iron non-adherence rate per 1000 population. This present research was approved by the institutional institutional review board.

### Statistical analysis

Continuous variables were displayed as either the mean and standard deviation or the median and interquartile range (IQR). Categorical variables were described as count (%). A paired t-test was used to compare EF readings at baseline and follow-up. The Wilcoxon signed-rank test was used to compare two related median values, as a normal distribution could not be assumed. The chi-square test was used to compare the primary outcome between the pre- and post-implementation groups. The Friedman test was employed to compare means of related groups for continuous variables. Missing values were imputed using a predictive mean matching multiple imputation model. If the Friedman test was significant, a Wilcoxon signed-rank test was used as the post hoc test for all paired comparisons, with the Bonferroni correction applied. All statistical analyses were performed using SPSS Version 29.0.2. GIS analysis and maps were created using ESRI ArcGIS Pro version 3.3.2 (2025).

## Results

### Baseline characteristics and iron deficiency consulting team performance

A total of 187 patients were included in the final cohort (Table 2). The majority of IV iron doses were for patients with HFrEF (53.5%). Eighty three % of iron deficiency was absolute iron deficiency, and more than 90% had transferrin saturation < 20%. Approximately 79% of patients received ferric carboxymaltose, and the remainder received iron sucrose. The median follow-up period of the IV iron consulting team was 372 (176, 623) days. The total IV iron induction dose (median (IQR)) was 1250 (750, 1250) mg. Fifty-seven patients (31.3%) were receiving oral iron therapy at baseline. Among 57 patients on oral iron therapy, the consulting team discontinued it in 19 patients (33.3%) during follow-up.

**Table 2.** Baseline characteristics of an iron deficiency cohort.

| Characteristics | Total (n=187) |
| --- | --- |
| Age (year) | 64.7 ± 15.4 |
| Gender (Male (%)) | 86 (46.0%) |
| Race (%) |  |
| White | 178 (95.2%) |
| Asian | 2 (1.1%) |
| Hispanic | 2 (1.1%) |
| Black | 5 (2.7%) |
| SBP (mm Hg) | 117.4 ± 17.1 |
| HR (beats/min) | 78.2 ± 15.2 |
| BMI (kg/m<sup>2</sup>) | 31.9 ± 8.0 |
| Weight (kg) | 92.1 ± 29.3 |
| Diabetes (%) | 106 (56.7%) |
| Hypertension (%) | 171 (91.4%) |
| Atrial fibrillation (%) | 108 (57.8%) |
| Previous stroke (%) | 40 (21.4%) |
| Dyslipidemia (%) | 156 (83.4%) |
| History of ASCVD (%) | 100 (53.5%) |
| Indication for intravenous iron (%) |  |
| HFrEF | 99 (53.5%) |
| HFpEF | 18 (9.7%) |
| PAH or CTEPH | 18 (9.7%) |
| PH (groups 2 and 3) | 9 (4.8%) |
| LVAD | 7 (3.8%) |
| HFmrEF | 6 (3.2%) |
| HFimpEF | 20 (10.8%) |
| Others | 9 (4.8%) |
| NYHA-FC<sup>#</sup> (%) | N=166 |
| 1 | 11 (6.6%) |
| 2 | 77 (46.4%) |
| 3 | 71 (42.8%) |
| 4 | 7 (4.2%) |
| ACC/AHA stages<sup>#</sup> (%) | N=160 |
| A | 0 |
| B | 1 (0.6%) |
| C | 147 (91.0%) |
| D | 12 (7.5%) |
| Serum creatinine (mg/dl) | 1.28 ± 0.96 |
| Potassium (mEq/L) | 4.28 ± 0.46 |
| Phosphorus (mg/dL) | 3.59 ± 0.67 |
| BNP (pg/mL) | 189.5 (73.5, 480.25) |
| Ferritin (mg/dL) | 44.0 (26.0, 80.6) |
| Iron saturation (%) | 12.2 ± 5.6 |
| Hemoglobin (g/dL) | 11.9 ± 1.9 |
| Absolute iron deficiency (%) | 156 (83.4%) |
| Transferrin saturation < 20% | 170 (90.9%) |
| Hb > 14 mg/dl | 29 (15.5%) |
| Left ventricle ejection fraction (%)^ | 34.3 ± 12.7% |
| Statin (%) | 133 (71.5%) |
| Beta-blocker (%)^ | 113 (85.0%) |
| ARNI (%)^ | 79 (59.8%) |
| MRA (%)^ | 102 (66.2%) |
| SGLT 2 I (%)^ | 80 (51.6%) |
| Loop use (%) | 140 (74.9%) |

ACC = American College of Cardiology; ACE-I = angiotensin converting enzyme inhibitor;
AHA = American Heart Association; ARB = angiotensin receptor blocker; ARNI = angiotensin neprilysin inhibitor; ASCVD = atherosclerotic cardiovascular disease; BMI = body mass index;
BNP = brain natriuretic peptide; CTEPH = chronic thromboembolic pulmonary hypertension;
HCM = hypertrophic cardiomyopathy; HFimpEF = heart failure with improved ejection fraction;
HFmrEF = heart failure with mildly reduced ejection fraction; HFpEF = heart failure with
preserved ejection fraction; HFrEF = heart failure with reduced ejection fraction; LVAD = left
ventricular assist device; MRA = mineralocorticoid receptor antagonist; NYHA FC = New York
Heart Association functional class; PAH = pulmonary arterial hypertension; SGLT 2 I = sodium-glucose cotransporter 2 inhibitor

### Outcomes

One hundred fifty-two patients (81.3%) were adherent to the appropriate use criteria, maintenance laboratory requirements, and dosing (Table 3). The most common reasons for nonadherence were the absence of a maintenance laboratory (15.5%), failure to administer all induction doses (1.6%), inappropriate use (1.1%), and incorrect dose (0.5%). Compared to the pre-implementation group, the primary outcome was significantly higher in the post-implementation group (81.3 vs. 40%, p<0.001).

**Table 3.** Outcome results of the 4-year experience cohort study.

|  | Pharmacist-led IV<br>iron clinic N=187 | Usual care N=42 | p-value |
| --- | --- | --- | --- |
| **Primary outcome:**<br>Adherence to the appropriate<br>criteria (use, dosing, and<br>laboratory requirements), n (%) | 152 (81.3%) | 17 (40.0%) | P<0.001 |
| Reasons for non-adherence: | 35 (18.7%) | N/A | N/A |
| No maintenance iron studies in<br>3-6 months after the induction | 29 (15.5%) |  |  |
| Incorrect dose | 1 (0.5%) |  |  |
| Inappropriate use | 2 (1.1%) |  |  |
| Not all induction doses given | 3 (1.6%) |  |  |
| Hospitalization for heart failure<br>within 1 year | 20 (10.9%) in 183<br>patients | 12 (28.4%) in 42<br>patients | 0.003 |
| Ejection fraction at 1-year<br>follow-ups | 42.3 ± 14.3% in 78<br>patients | N/A | N/A |
| 1-year mortality | 11 (6%) in 184<br>patients | 3 (7.1%) in 42<br>patients | 0.778 |
| Adverse drug effects (%) | 10 (5.3%) | N/A | N/A |

Displayed as count (%) or mean ± standard deviation

Hospitalization for HF at one year was 10.9% and one-year all-cause mortality was 6.0%. The median left ventricular EF at the follow-up was 43.9 (32.5, 55.0) % (p<0.001 compared with the median EF 35.0 (25.0, 44.0) % at baseline) (Table 4).

**Table 4.**
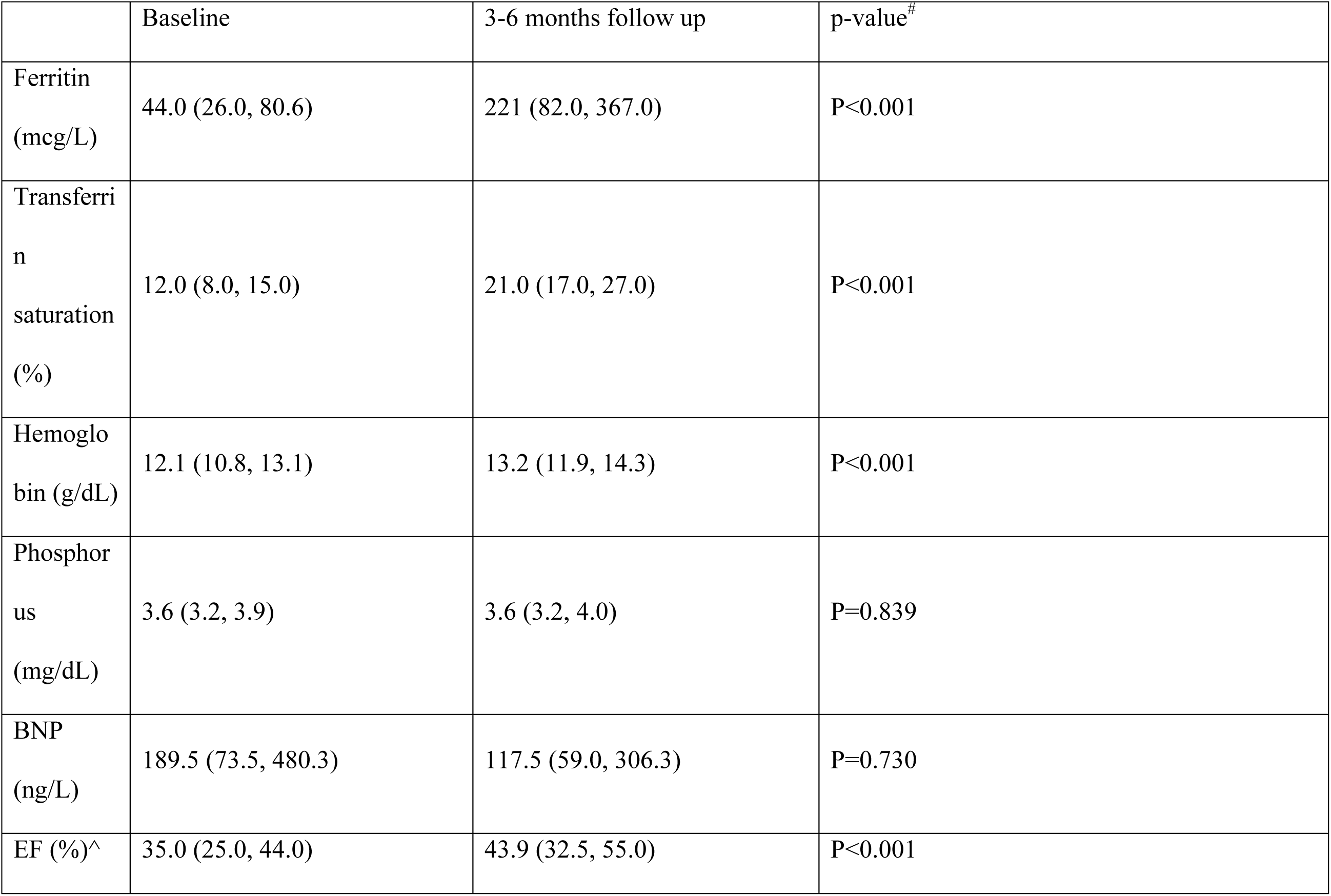
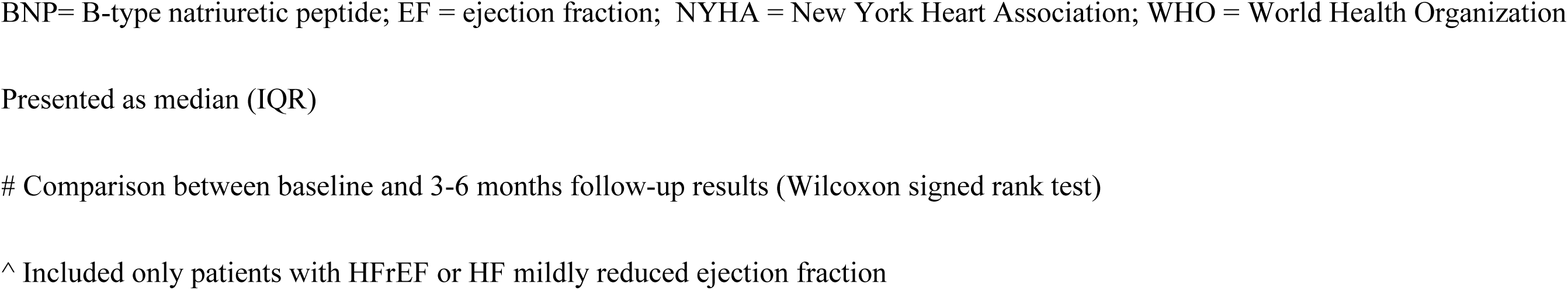
Clinical secondary outcomes between baseline and 3-6 month follow-up after induction course.

|  | Baseline | 3-6 months follow up | p-value<sup>#</sup> |
| --- | --- | --- | --- |
| Ferritin<br>(mcg/L) | 44.0 (26.0, 80.6) | 221 (82.0, 367.0) | P<0.001 |
| Transferrin<br>saturation<br>(%) | 12.0 (8.0, 15.0) | 21.0 (17.0, 27.0) | P<0.001 |
| Hemoglobin<br>(g/dL) | 12.1 (10.8, 13.1) | 13.2 (11.9, 14.3) | P<0.001 |
| Phosphorus<br>(mg/dL) | 3.6 (3.2, 3.9) | 3.6 (3.2, 4.0) | P=0.839 |
| BNP<br>(ng/L) | 189.5 (73.5, 480.3) | 117.5 (59.0, 306.3) | P=0.730 |
| EF (%)^ | 35.0 (25.0, 44.0) | 43.9 (32.5, 55.0) | P<0.001 |

BNP= B-type natriuretic peptide; EF = ejection fraction; NYHA = New York Heart Association; WHO = World Health Organization
Presented as median (IQR)

### Long-term iron therapy outcomes

Ferritin, iron saturation, and hemoglobin values at cycle one were significantly increased after an IV iron induction course compared with baseline values and were numerically maintained throughout follow-up (Table 5). Phosphorus levels remained within the normal range throughout the follow-up period.

**Table 5.** Longitudinal changes in laboratory results, treatment achievement goals, and oral iron therapy over four years.

|  | Baseline | Cycle 1 | Cycle 2 | Cycle3 | Cycle 4 | Cycle 5 | Cycle 6 | Cycle 7 | p-value |
| --- | --- | --- | --- | --- | --- | --- | --- | --- | --- |
| Ferritin (mcg/L) | 44.0 (26.0, 80.6)<br>(n=187) | 221.0 (82.0, 367.0)<br>(n=159) | 153.0 (83.0, 292.0)<br>(n=115) | 138.5 (67.3, 287.0)<br>(n=72) | 180.5 (85.0, 329.5)<br>(n=40) | 189.0 (83.0, 322.5)<br>(n=25) | 184.5 (119.3, 268.8)<br>(n=12) | 136.0 (86.0, 348.5)<br>(n=9) | P<0.001 |
| Transferrin saturation (%) | 12.0 (8.0, 15.0)<br>(n=187) | 21.0 (17.0, 27.0)<br>(n=159) | 21.0 (16.0, 28.0)<br>(n=115) | 19.0 (14.0, 25.0)<br>(n=72) | 21.0 (15.0, 27.0)<br>(n=42) | 20.0 (17.0, 25.0)<br>(n=25) | 17.0 (14.0, 20.8)<br>(n=12) | 18.0 (16.5, 29.5)<br>(n=9) | P<0.001 |
| Hemoglobin (g/dL) | 12.1 (10.8, 13.1)<br>(n=187) | 13.2 (11.9, 14.3)<br>(n=162) | 13.2 (12.1, 14.1)<br>(n=113) | 12.7 (11.6, 14.1)<br>(n=71) | 12.5 (11.8, 13.7)<br>(n=42) | 12.5 (11.7, 13.6)<br>(n=23) | 12.9 (11.7, 13.9)<br>(n=12) | 13.1 (11.9, 13.9)<br>(n=8) | P<0.001 |

Abbreviation: BNP = B-type natriuretic peptide, ID = iron deficiency, IV = intravenous
Each cycle is approximately 6 months
Presented as median (IQR)
P-value was after multiple imputation and Bonferroni adjustment
Friedman test and if the Friedman test was significant, a Wilcoxon signed rank test was used as a post-hoc test for all paired comparisons with Bonferroni correction. \* p<0.05 compared with the baseline level

### Geospatial analysis

Geospatial analysis showed denser patient clusters around infusion centers in the density heat map, represented as yellow-to-red areas (Figure 1). The driving time to the closest infusion centers was significantly shorter than to the Ruby infusion center (26.3±13.4 vs. 89.6±53.2 minutes, p<0.001). Drive times for all patients were within 75 minutes, with 49% being within 15 minutes of an infusion center, while up to 5 hours of drive time to Ruby and only 8% being within 15 minutes were required. Geospatial analysis of the bivariate map showed that higher areas of guidance non-adherence (i.e., the dark purple and dark blue areas on the maps) occurred only at greater drive times farther from infusion centers (Figure 2).

**Figure 1.**
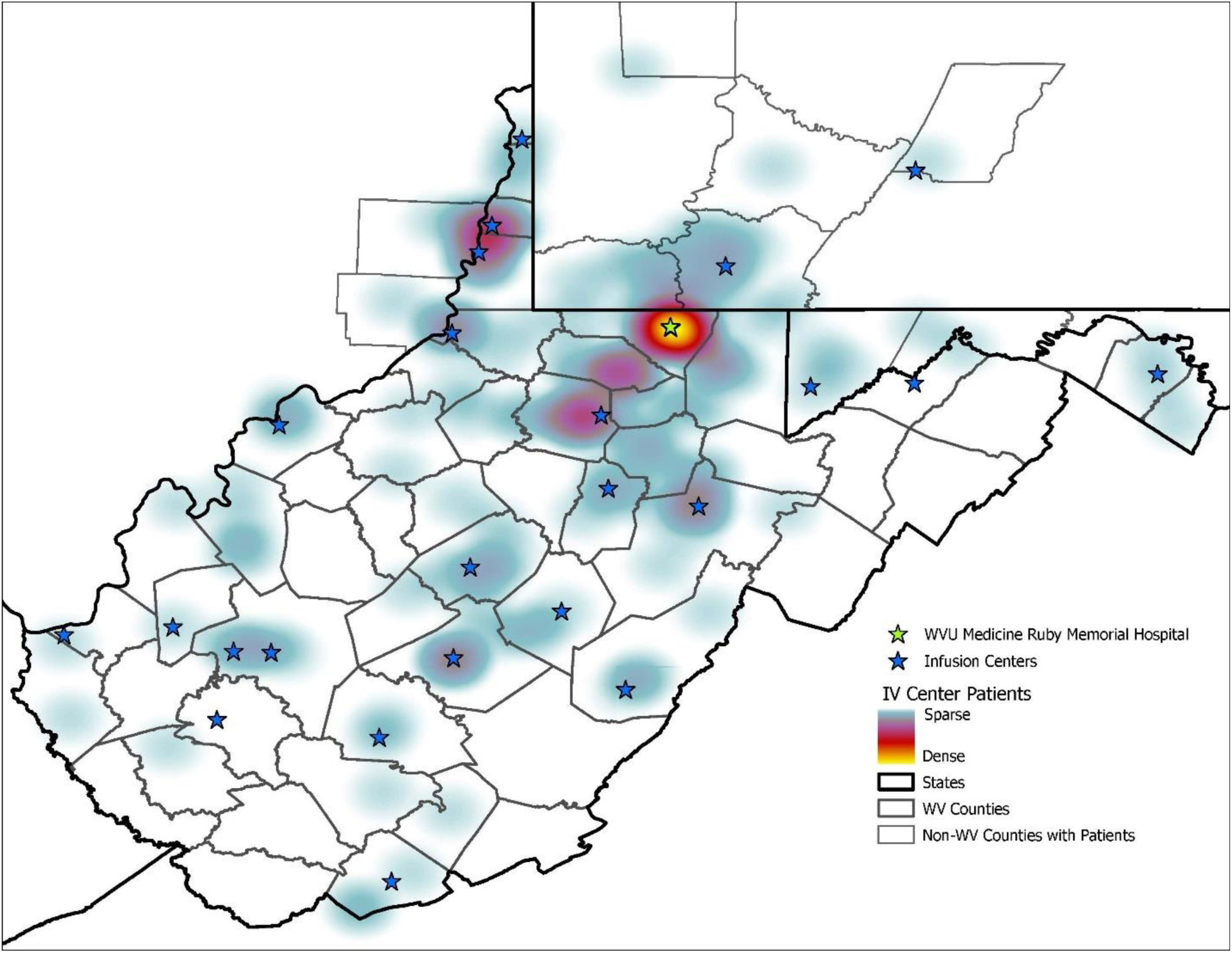
The heat map describing the density of patients around areas in IV iron infusion sites

**Figure 2.**
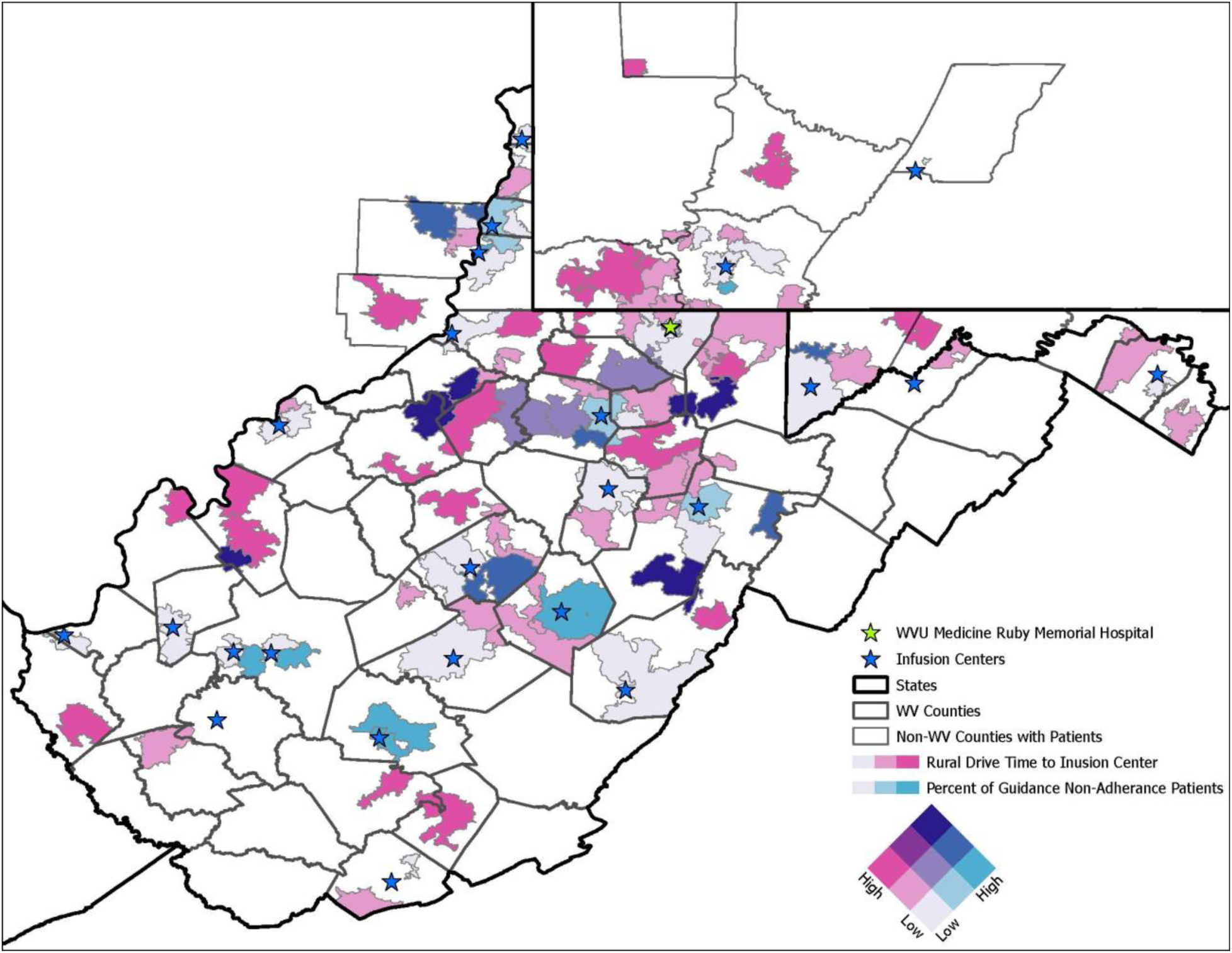
The bivariate map comparing drive time to an infusion center versus the IV iron non-adherence patient percentage

## Discussion

A pharmacists-providers collaborative iron deficiency clinic coordinated IV iron therapy for 187 patients. Among 57 patients on oral iron therapy, the consulting team discontinued it in about one-third during follow-up. It significantly improved adherence rate to the IV iron appropriate use criteria, laboratory requirements, and dosing compared to the pre-implementation group.

The pharmacists-providers collaborative iron deficiency clinic achieved an adherence rate of approximately 80% to the IV iron guidance. About 82% of non-adherences was due to no follow-up labs within 6 months of induction therapy. These patients either lost follow-ups with the HF service or obtained labs more than 6 months after induction therapy. All non-adherent patients were contacted by the iron deficiency clinic at least three times before they were determined to be nonadherent for the follow-up labs. A potential solution to improve this adherence rate is to reinforce the importance of repeating iron studies and hemoglobin labs to promptly assess the needs for IV iron maintenance doses.

The adherence rate in the present study was numerically lower than that in the previously reported pilot study results (81 % vs. 93%), although both were significantly higher than the pre-implementation group (40%).^10^ The numerical differences in the adherence rate between the two different studies are likely attributed to changes in the number of patients, ranging from 81 to 187 patients, or access to laboratories in rural areas. Over the last few years, our iron deficiency clinic has handled a greater number of IV iron cases across the Appalachian region, particularly in rural areas. The numerical decrease in the adherence rate may have reflected more challenging cases for IV iron therapies, which are more likely to have had access issues with IV iron infusion centers. Our geospatial analysis results demonstrated that higher areas of guidance non-adherence occurred only at greater drive times farther from infusion centers. Thus, the solution to improve adherence is to increase the number of IV iron infusion centers across the Appalachian region and to improve access to IV iron therapy by shortening drive times to medical facilities that can offer IV iron infusions and complete laboratory requirements for IV iron therapy.

Limited guidance is available on the duration and monitoring plan for long-term IV iron therapy in practice. HF pharmacists follow up on the labs every 6 months, and approximately 400-600 mg of IV iron therapy is administered whenever patients meet the criteria for iron deficiency. The median follow-up period was approximately one year, and only a limited number of patients had follow-up longer than two years. Further investigations are needed to evaluate the frequency of labs and maintenance labs (every three vs. every six months) and the appropriate criteria for maintenance doses (HF guideline-based criteria vs. other criteria, such as iron saturation-based criteria).

The present study showed that 31.3 % of patients in the study were taking oral iron therapy. The IRON-OUT trial failed to show the benefits of oral iron therapy in patients with HF.^13^ Since it is known that polypharmacy and pill burdens affect adherence to pharmacotherapy, ideally, all oral iron therapy can be discontinued once IV iron therapy is initiated.^14^ Our study showed that oral iron therapy was discontinued in approximately one third of patients over a median follow-up of one year, while the remaining of two thirds was continued on oral iron therapy. This persistence more likely attributes to clinical inertia and the fact that stopping oral iron therapy was not incorporated with the formal workflow of intravenous iron therapy service. Deprescribing oral iron therapy and reducing pill burdens are the advantages of IV iron therapy. We, therefore, propose that deprescribing oral iron therapy should be an explicit criterion for initiating IV iron therapy.

### Limitations

This is a retrospective cohort study, and no causal inference can be made. The most common reason for nonadherence was the absence of follow-up labs within 6 months of the induction course. However, the study lacked sufficient information about the actual timing of all follow-up labs after the induction course. Next, the main results of the present study are only applicable to patients receiving ferric carboxymaltose, as approximately 80% of the patients in the study received it. Further investigation is needed to compare outcomes between ferric carboxymaltose and iron sucrose to apply this data to patients receiving iron sucrose. Third, our study used the adherence to the appropriate IV iron use, dosing and monitoring as a primary outcome. Clinical outcomes such as hospitalization for HF were evaluated as secondary outcomes. Clinical outcomes as primary outcomes can be considered in future larger scale prospective studies.

## Conclusion

The pharmacists-providers collaborative IV iron clinic significantly improved the performance of IV iron therapy in rural heart failure care settings compared to the usual care. Prospective studies are needed to investigate the effectiveness of the pharmacists-providers collaborative IV iron clinic.

## Data Availability

The availability of all data is upon request to the corresponding author.

## Figure legends

Central Illustration. Four-year performance of the Pharmacists-providers collaborative iron deficiency clinic

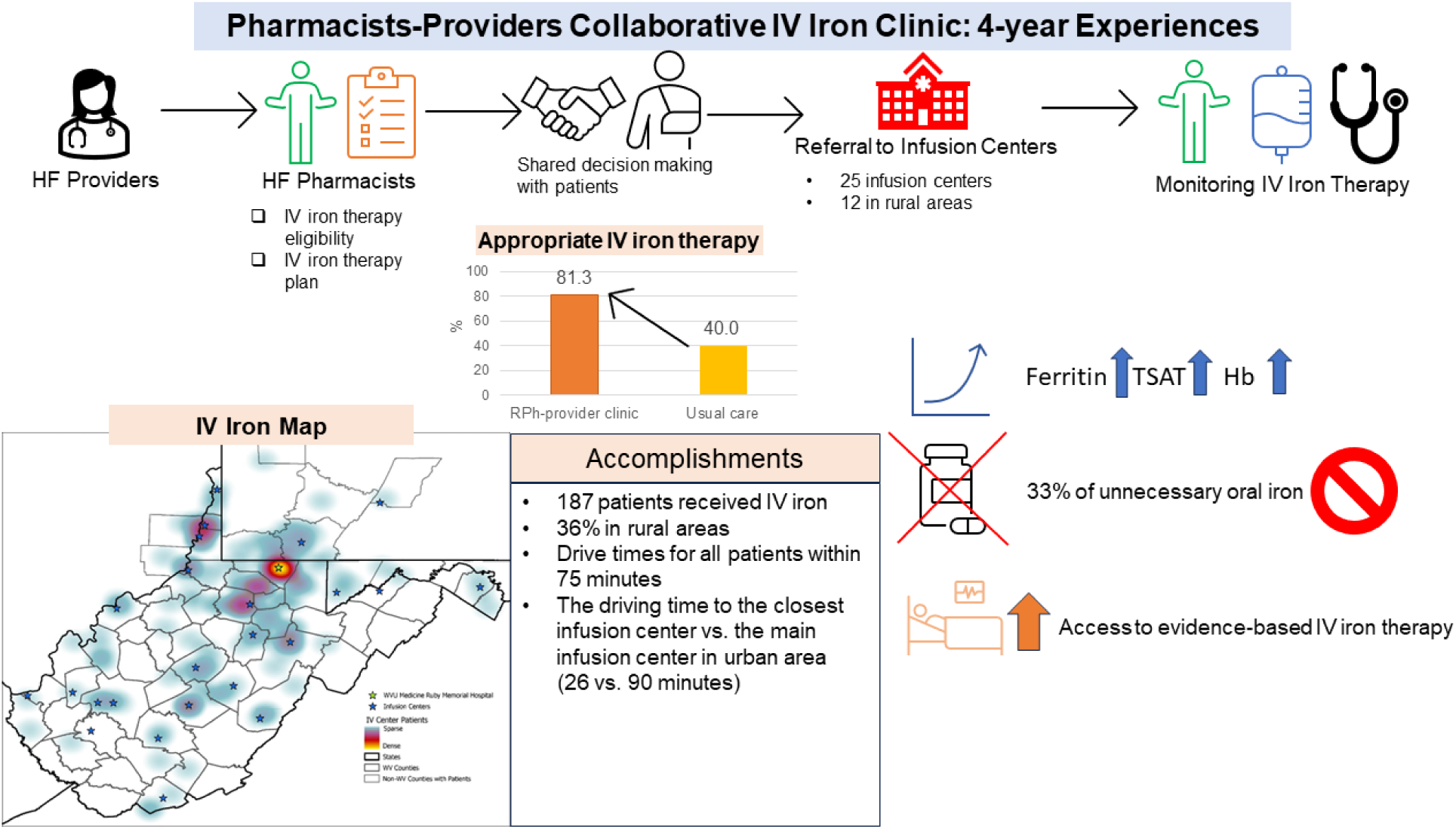

Abbreviations: Hb = hemoglobin; HF = heart failure; IV = intravenous; TSAT = transferrin saturation

## Conflict of Interests Statement

Kazuhiko Kido is an independent contractor as a topic editor for DynaMed, LLC^®^.

## Acknowledgement

Research reported in this publication was supported by the National Institute of General Medical Sciences of the National Institutes of Health under Award Number 5U54GM104942-08. The content is solely the responsibility of the authors and does not necessarily represent the official views of the National Institutes of Health.

## Notes

### Competing Interest Statement

The authors have declared no competing interest.

### Author Declarations

West Virginia University IRB provided approval for the research.

